# Metformin use is associated with lower mortality from bacterial sepsis and improved immunocompetence in Thai diabetes patients with acute melioidosis

**DOI:** 10.1101/2025.10.29.25338967

**Authors:** Barbara Kronsteiner, Clement Twumasi, Priyanka Abraham, Suchintana Chumseng, Panjaporn Chaichana, Phumrapee Boonklang, Arin Wongprommoon, Kesorn Angchagun, Narisara Chantratita, Direk Limmathurotsakul, Nicholas P.J. Day, Parinya Chamnan, Claire Chewapreecha, Susanna J. Dunachie

**Affiliations:** Peter Medawar Building for Pathogen Research, Nuffield Dept. of Medicine, University of Oxford, Oxford, UK; NDM Centre for Global Health Research, Nuffield Dept. of Medicine, University of Oxford, Oxford, UK; NIHR Oxford Biomedical Research Centre, Oxford University Hospitals NHS Foundation Trust, Oxford, UK; Mahidol Oxford Tropical Medicine Research Unit, Faculty of Tropical Medicine, Mahidol University, Bangkok, Thailand; Department of Microbiology and Immunology, Faculty of Tropical Medicine, Mahidol University, Bangkok, Thailand; Department of Tropical Hygiene, Faculty of Tropical Medicine, Mahidol University, Bangkok, Thailand; Centre for Tropical Medicine and Global Health, Nuffield Department of Medicine, University of Oxford, UK; Cardiometabolic Research Group, Sunpasitthiprasong Hospital, Ubon Ratchathani, Thailand

**Keywords:** Metformin, Diabetes, Sepsis, Melioidosis, Protection

## Abstract

**Background:** Diabetes mellitus (DM) is a major risk factor for acquiring infections. Metformin, the first-line treatment for type 2 DM, is associated with beneficial outcome from various infectious diseases. The neglected tropical disease melioidosis has up to 50% in-hospital mortality rate and the highest risk association of DM with any infectious disease (12-fold increased risk). A better understanding of the impact of anti-hyperglycaemic drug treatment on disease outcome is needed.

**Methods:** We analysed the association of anti-hyperglycaemic treatment on outcome from acute melioidosis in 273 Thai patients with known DM using logistic regression and classification tree modelling while controlling for age, sex, HbA1c and history of renal impairment. Immunological parameters including T-cell and humoral responses to the pathogen as well as cytokine levels in blood were measured in a subset of individuals.

**Results:** We report higher survival rates in acute melioidosis patients with DM who were taking the glucose-lowering drug metformin prior to infection. Metformin use was associated with a 50% decrease in 28-day mortality whilst renal impairment was associated with a doubling of mortality after adjusting for covariates. Unsupervised classification tree modelling further identified glycaemic control and age as modifying factors of outcome. Furthermore, metformin treatment was associated with greater T-cell responsiveness and the presence of T-cell modulating cytokines.

**Conclusions:** We provide evidence for a protective effect of metformin in melioidosis and identify HbA1c and age as additional clinically relevant predictors of outcome. Further mechanistic research and randomized controlled trials are required to explore a causal link between metformin and protection from infectious diseases.

**Highlights:**

- Metformin treatment in diabetes is associated with reduced risk of death from acute melioidosis
- Metformin has a beneficial impact on immunological parameters in acute melioidosis
- Machine learning identifies glycaemic control and age as modifying factors of outcome in diabetes

## INTRODUCTION

Diabetes mellitus (DM) is a major risk factor for acquiring infections including tuberculosis, melioidosis, influenza and dengue [1]. The recent COVID-19 pandemic has underscored the susceptibility of people with DM to infection and more severe outcomes [2] and highlighted the need for better understanding of immune responses and correlates of protection in this patient group. The prevalence of DM worldwide is rapidly increasing with a high burden seen in low- and middle-income countries [1]. The WHO recommends metformin as first-line treatment for type 2 diabetes mellitus (T2DM) [3], with 89% of people living with T2DM in the UK having received metformin as initiation prescription in 2017 [4]. However, many of the world’s population living with T2DM are not taking it due to various reasons including side effects, being prescribed an alternative medicine from the start or metformin not being continued when treatment is intensified by adding other anti-hyperglycaemic medication [5]. Metformin has received much recent attention as a host-directed therapy across different research fields including ageing [6] and cancer [7]. There is a growing body of evidence that oral metformin treatment improves outcomes from various infectious diseases including sepsis [8], TB [9], COVID-19 [10] and hospital-acquired infection [11].

Melioidosis is a neglected tropical disease caused by the intracellular bacterium *Burkholderia pseudomallei* (BP), with a case fatality rate exceeding 40% in Northeast Thailand [12]. The disease disproportionately affects people with DM, with over 50% of acute melioidosis patients having DM [13]. Other risk factors include chronic kidney and lung disease, hazardous alcohol consumption and immunosuppressive therapies [14]. A survival benefit was previously demonstrated in acute melioidosis patients taking the glucose-lowering sulphonylurea drug glyburide [15]. A prospective cohort study determining factors of all-cause mortality in over 9,300 Thai diabetes patients showed an all-cause survival benefit in patients taking metformin reducing the risk of death by 40% [16], after adjustment for patient-related factors.

The impact of metformin treatment on outcome from melioidosis is currently unknown. Using three prospective longitudinal cohort studies of acute melioidosis conducted in Northeast Thailand we show that metformin is associated with favourable survival outcomes from melioidosis.

## METHODS

### Study design and participants

We analysed the association of metformin treatment on outcome in three prospective longitudinal cohort studies of acute melioidosis between 2012 and 2022 at Sunpasitthiprasong Hospital in Ubon Ratchatani, Thailand[17]. Patients who presented to the hospital with acute illness were enrolled upon culture-confirmed diagnosis of melioidosis [18]. DM status was evaluated by HbA1c measurement in all enrolled patients, and a new diagnosis of DM was made if HbA1c was >6.5%, according to WHO criteria [19]. Study inclusion criteria for this analysis were pre-existing or newly diagnosed DM on admission to hospital. The exclusion criteria were lack of outcome data, incomplete data on anti-hyperglycaemic medication or reported as not taking medication as prescribed or those who were diet controlled. Medications included metformin, sulphonylureas (chiefly glipizide), insulin, pioglitazone and sitagliptin. No formal typing of DM was available but the vast majority of people in our cohort are likely to have type 2 diabetes due to typical presentations being insidious and not requiring insulin from the start, alongside current knowledge of DM types in the region. Anyone diagnosed with DM during study enrolment was categorised as newly diagnosed. The primary outcome was death within 28-days of enrolment (28-day mortality). Study protocols were approved by the ethics committees of: the Faculty of Tropical Medicine, Mahidol University (MUTM 2012-018-01, MUTM 2015-071-01); Sunpasitthiprasong Hospital, Ubon Ratchathani (018/2555, 017/2559, 015/62C); and the Oxford Tropical Research Ethics Committee (OxTREC 519-19, 520-19 and 25-19). Studies were conducted according to the principles of the Declaration of Helsinki (2008) and the International Conference on Harmonization (ICH) Good Clinical Practice (GCP) guidelines. Written informed consent was obtained for all participants enrolled in the study.

### Immunological parameters

Peripheral blood mononuclear cells (PBMC) were isolated from whole blood and analysed for IFN-γ secretion in response to heat-inactivated (HIA) BP antigens using an enzyme-linked immunospot assay (ELISpot) [18]. Serological responses to BP were determined by indirect hemagglutination assay (IHA) [20]. Protein levels in serum were measured using the Mesoscale Discovery (MSD) V-PLEX Human Biomarker 54-Plex Kit to measure the concentrations of 54 analytes of interest (**Supplementary Table 1**). The assay was performed as per manufacturer’s instructions and the plates were measured immediately on the MSD Sector QuickPlex SQ 120MM Reader. Details of data analysis are provided in the **Supplementary information**.

### Statistical Analysis

Categorical variables were expressed as counts and frequencies and compared using Fisher’s exact test. Continuous variables were expressed as mean and range for parametric data (age) and differences between groups were calculated using Student’s *t*-test. Non-parametric variables (immunological parameters, HbA1c) were expressed as median and interquartile range and differences between groups were calculated using the Mann-Whitney U test. To evaluate the relationship between anti-hyperglycaemic treatment variables and 28-day mortality among patients with acute melioidosis multivariable logistic regression models (to estimate odds ratios), and unsupervised classification tree models (to explore the hierarchical structure of predictive factors) were performed in R. The effect of metformin on cytokine profiles in a subgroup of patients was explored using permutation-based Hotelling’s T2 test followed by Lasso-regularised multivariate regression. More details are provided in the **Supplementary information**.

## RESULTS

### Patient Characteristics

Out of the 557 patients diagnosed with acute melioidosis and DM co-morbidity, 3 were excluded due to missing data on 28-day mortality and 201 were excluded because they reported not taking any anti-hyperglycaemic medication (including those on diet control) or where records were inconclusive. Of the remaining 353 patients, 75 were newly diagnosed with DM upon hospital admission and therefore excluded from analysis. A total of 278 patients with pre-existing DM were included in the study cohort (**Figure 1A**). Among those 118 patients were taking metformin, 108 patients were taking sulphonylureas (chiefly glipizide) and 186 patients were taking insulin. There was substantial overlap in treatments with many patients taking multiple oral anti-hyperglycaemic drugs with or without insulin (**Figure 1B**). A small number of individuals (n=11) were taking pioglitazone in various combinations with other oral drugs or insulin and one patient was taking sitagliptin. Next, we split the cohort into two groups, patients who survived and patients who died within 28-days of hospital admission (**Table 1**).

**Figure 1.**
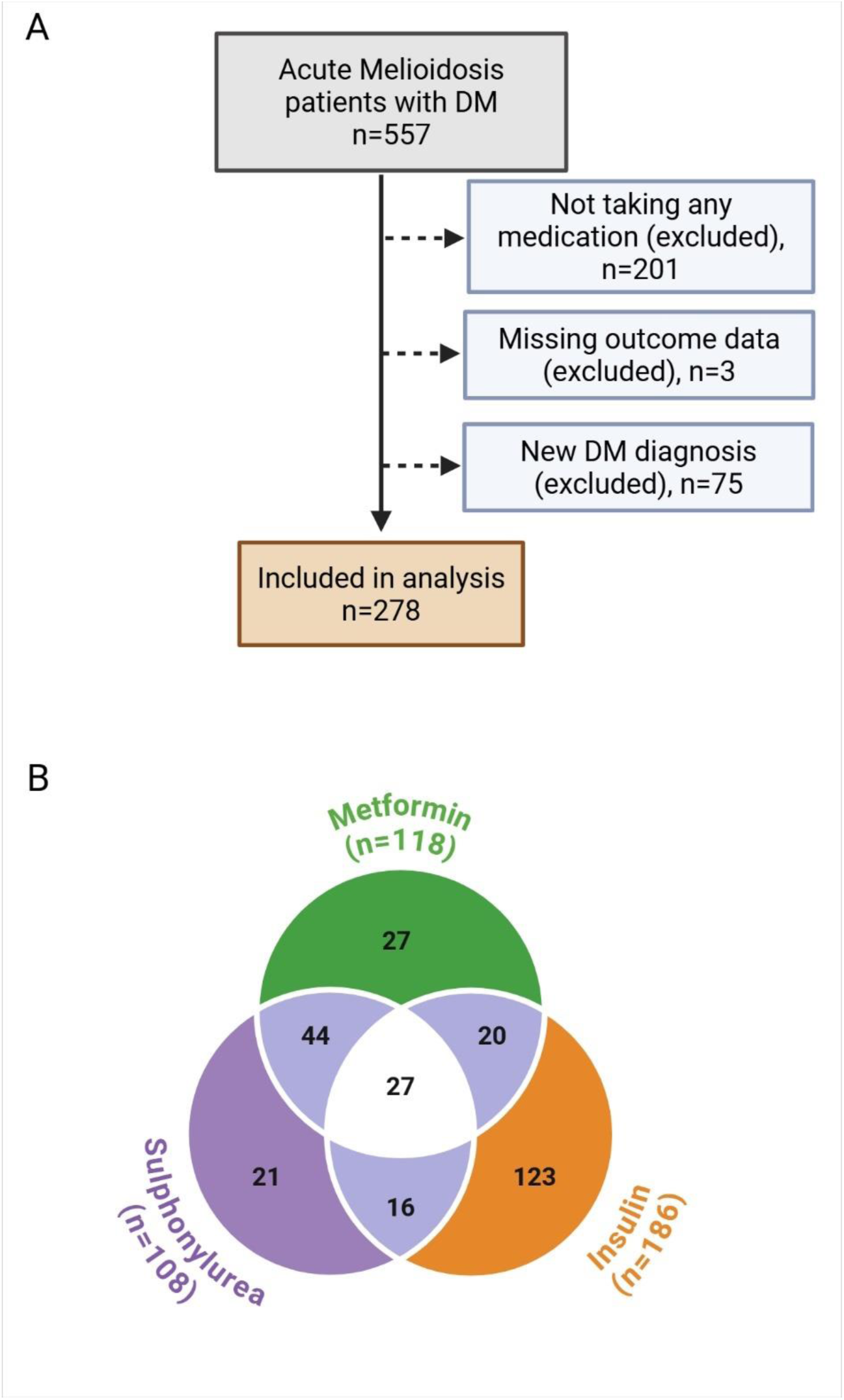
Inclusion criteria and anti-hyperglycaemic drug treatment. (A) Flow chart of inclusion criteria and (B) glucose lowering drug treatments in 278 patients with known diabetes and acute melioidosis. Numbers in Venn diagram indicate the number of patients taking the respective drug combination. The numbers in brackets indicate the total number of patients on the respective drug (including those on multiple treatments).

**Table 1.** Acute melioidosis patient characteristics by outcome.

| Characteristics | Survived<br>n=209 | Died*<br>n=69 | p value^ |
| --- | --- | --- | --- |
| **Sex** |  |  | 0.959 |
| Female, n | 75 | 25 |  |
| Male, n | 134 | 44 |  |
| **Age, mean in years (min-max)** | 52 (19-89) | 54 (26-80) | 0.144 |
| **History of renal impairment, n (%)** | 50 (23.9) | 31 (44.9) | <0.001 |
| **HbA1C, median in % (IQR)<sup>+</sup>** | 10.4 (8.5-12.6) | 11.0 (7.9-13.3) | 0.732 |
| **Anti-hyperglycaemic treatment<sup>#</sup>** |  |  |  |
| Insulin, n (%) | 132 (53.9%) | 54 (63.5%) | 0.021 |
| Metformin, n (%) | 101 (41.2%) | 17 (20%) | <0.001 |
| Sulphonylurea, n (%) | 91 (37.1%) | 17 (20%) | 0.005 |

\*28-day mortality, ^Mann Whitney U test (HbA1C), t-test (age) and Pearson's Chi-square test (categorical data), <sup>+</sup>missing data for n=5, <sup>#</sup>prior to hospital admission

Among the 278 patients with acute melioidosis, 25% (n=69) died. Sex distribution, age and HbA1c were similar between those who died and survived. Among those who died compared to those who survived, the proportion of patients with a history of renal impairment (died: 42.4 % vs survived: 20.8%, p<0.001) was significantly higher. The proportion of patients with pre-existing DM who were taking metformin or a sulphonylurea drug was significantly higher in survivors compared to those who died (metformin: survived 41.2%, died 20%, p<0.001; sulphonylurea: survived 37.1%, died 20%, p=0.005), whereas insulin use was more common in individuals who died (survived 54.9%, died 63.5%, p=0.021).

### Metformin treatment and absence of renal impairment are associated with better outcome

We used logistic regression analysis in order to quantify the strength and direction of association of anti-hyperglycaemic drug treatment in patients with a DM diagnosis prior to hospital admission with mortality outcomes. A total of 273 cases were included in the analysis after removing 5 cases due to HbA1c data missing at random (see Supplementary Information, Methods). We evaluated two models, one including metformin and sulphonylurea use as distinct binary predictors (separate oral drugs) and a second one combining oral drug treatment into one single binary indicator (combined oral drugs), while adjusting for age, sex, history of renal impairment, insulin use and HbA1c. Using a stepwise regression approach the final models included sex, history of renal impairment and HbA1c as covariates, whereas the inclusion of insulin treatment and age as additional predictors did not result in the best-fitted regression models and were therefore omitted.

Among patients with acute melioidosis, logistic regression analyses (**Table 2**) identified renal failure and oral anti-hyperglycaemic drug use as significant predictors of 28-day mortality across both modelling scenarios. In the best-fitted logistic regression model (with prediction accuracy of 76.9% and AUC value of 0.754) where metformin and sulphonylurea use were included as separate predictors, the presence of renal failure was associated with nearly two-fold increased odds of mortality (adjusted OR = 1.95, 95% CI: 1.02–3.70; *p* = 0.042), while metformin use was associated with over 50% reduced odds of mortality (adjusted OR = 0.46, 95% CI: 0.22–0.94; *p* = 0.033). Sulphonylurea use, although suggestive of a protective effect, did not reach statistical significance (adjusted OR = 0.62, 95% CI: 0.31–1.26; *p* = 0.189). In the alternative model (with prediction accuracy of 80.8% and AUC value of 0.688) using a single combined oral drug binary indicator, oral anti-hyperglycaemic treatment remained a strong and statistically significant protective factor associated with nearly 70% reduction in the likelihood of mortality (adjusted OR=0.31, 95% CI: 0.17–0.59; *p* < 0.001). Renal failure continued to be associated with significantly increased odds of mortality (adjusted OR = 1.97, 95% CI: 1.03–3.76; *p* = 0.039). Across both models, neither sex nor HbA1c levels were statistically significant predictors after model adjustment.

**Table 2.** Estimated crude and adjusted odds ratios (OR) of significant predictors of mortality among patients with acute melioidosis under the two modelling scenarios based on the best-fitted logistic regression models.

| Predictors | Crude OR | Adjusted OR | 95% CI | p-value |
| --- | --- | --- | --- | --- |
| **Logistic Regression Model with separate oral anti-hyperglycaemic drugs** |  |  |  |  |
| Sex: male vs female | 1.09 | 1.12 | (0.59, 2.11) | 0.734 |
| History of renal impairment: yes vs no | 2.49 | 1.95 | (1.02, 3.70) | 0.042* |
| HbA1c (%) (continuous) | 0.95 | 0.96 | (0.88, 1.06) | 0.458 |
| Drug: metformin vs no metformin | 0.34 | 0.46 | (0.22, 0.94) | 0.033* |
| Drug: sulphonylurea vs no sulphonylurea | 0.45 | 0.62 | (0.31, 1.26) | 0.189 |
| **Logistic Regression Model with combined oral anti-hyperglycaemic drug indicator** |  |  |  |  |
| Sex: male vs female | 1.09 | 1.17 | (0.61, 2.21) | 0.638 |
| History of renal impairment: yes vs no | 2.49 | 1.97 | (1.03, 3.76) | 0.039* |
| HbA1c (%) (continuous) | 0.95 | 0.96 | (0.87, 1.06) | 0.422 |
| Drug: oral drug vs no oral drug | 0.29 | 0.31 | (0.17, 0.59) | < 0.001*** |

\*\*\*p<0.001, \*\*p<0.01, \*p<0.05. CI denotes Confidence Interval. Insulin use and age were excluded as additional predictors in the logistic regression models. The models were fitted based on training dataset (90%, n=247 patients) and cross-validated based on the testing set (10%, n=26 patients).

**Table 3.** Estimated crude and adjusted odds ratios (OR) of significant predictors of BP specific T cell and antibody responsiveness in acute melioidosis patients with DM.

| Predictors | Crude | Adjusted |  |  |
| --- | --- | --- | --- | --- |
|  | OR | OR | 95% CI | p-value |
| **Logistic regression model: T cell responders\*** |  |  |  |  |
| Age (ys, continuous) | 1.00 | 1.00 | (0.97, 1.04) | 0.873 |
| Sex: male vs female | 0.57 | 0.51 | (0.22, 1.19) | 0.119 |
| 28-day Mortality: yes vs no | 0.42 | 0.48 | (0.19, 1.23) | 0.124 |
| History of renal impairment: yes vs no | 2.38 | 2.98 | (0.97, 9.15) | 0.057 |
| HbA1c (% continuous) | 1.02 | 1.08 | (0.92, 1.28) | 0.364 |
| Drug: insulin vs no insulin | 0.30 | 0.29 | (0.07, 1.19) | 0.085 |
| Drug: metformin vs no metformin | 2.93 | 3.52 | (1.15, 10.80) | **0.028** |
| Drug: sulphonylurea vs no sulphonylurea | 1.45 | 0.59 | (0.18, 1.91) | 0.380 |
| **Logistic regression model: Antibody responders^** |  |  |  |  |
| Age (ys, continuous) | 0.98 | 0.99 | (0.94, 1.03) | 0.482 |
| Sex: male vs female | 0.75 | 0.77 | (0.31, 1.95) | 0.580 |
| 28-day Mortality: yes vs no | 0.33 | 0.29 | (0.11, 0.78) | **0.014** |
| History of renal impairment: yes vs no | 0.52 | 0.45 | (0.16, 1.28) | 0.133 |
| HbA1c (% continuous) | 0.99 | 0.95 | (0.80, 1.12) | 0.532 |
| Drug: insulin vs no insulin | 0.54 | 0.43 | (0.11, 1.73) | 0.235 |
| Drug: metformin vs no metformin | 1.17 | 0.85 | (0.27, 2.69) | 0.786 |
| Drug: sulphonylurea vs no sulphonylurea | 0.86 | 0.45 | (0.14, 1.48) | 0.188 |

\*n=121 cases, ^n=127 cases included in analysis, CI: confidence interval

### Machine learning identifies glycaemic control and age as additional modifying factors for outcome from infection

We next used an unsupervised machine learning approach employing a classification tree model to predict 28-day mortality among patients with acute melioidosis. As before we used a train dataset (90%, n=247) and two scenarios: separate oral drugs and combined oral drugs alongside other covariates (sex, age, renal failure status, HbA1c, and insulin use). The tree illustrates how combinations of treatment, clinical and demographic variables relate to mortality outcome in an unsupervised manner, with cut-offs determined by the model and reflecting the specific structure of the dataset used.

The model using separate anti-hyperglycaemic drugs (accuracy 80.8%, AUC 0.767, **Figure 2A**) shows that metformin use is the most influential split (p<0.05) with patients on metformin (41% of the total training data cohort) being more likely to survive (86% Alive), and further stratification based on HbA1c levels and sulphonylurea treatment. Among metformin users with a higher HbA1c (≥ 6.4%), mortality was low (11%), whereas mortality was notably higher (60%) in those with lower HbA1c (< 6.4%). Notably, the latter group only represent 2% of the entire cohort. For patients not taking metformin (59% of the total training cohort), sulphonylurea use further differentiated outcomes. Those taking sulphonylurea had a low mortality rate (18%) irrespective of their age. Outcome in patients not on sulphonylurea was further modified by age with patients younger than 71 years showing intermediate mortality rates (42%), while older patients (≥ 71 years) had a high mortality rate (71%). The second model using a combined binary predictor of oral anti-hyperglycaemic drug use (accuracy 76.9%, AUC 0.658, **Figure 2B**) shows that the use of metformin and/or sulphonylurea is the most influential predictor (p<0.05) of outcome with patients taking any oral drug (56% of the training data cohort) being more likely to survive (86% Alive). Similar to the first model, glycaemic control further discriminated outcome in patients taking oral drugs and age was a modifying factor in patients on insulin (not taking any oral drugs) with lower mortality rate in patients with HbA1c ≥ 6 and being under 71ys respectively. Importantly, the individuals at highest risk of dying only represent a small proportion of the entire cohort (2-3%), reflecting the most vulnerable cases.

**Figure 2.**
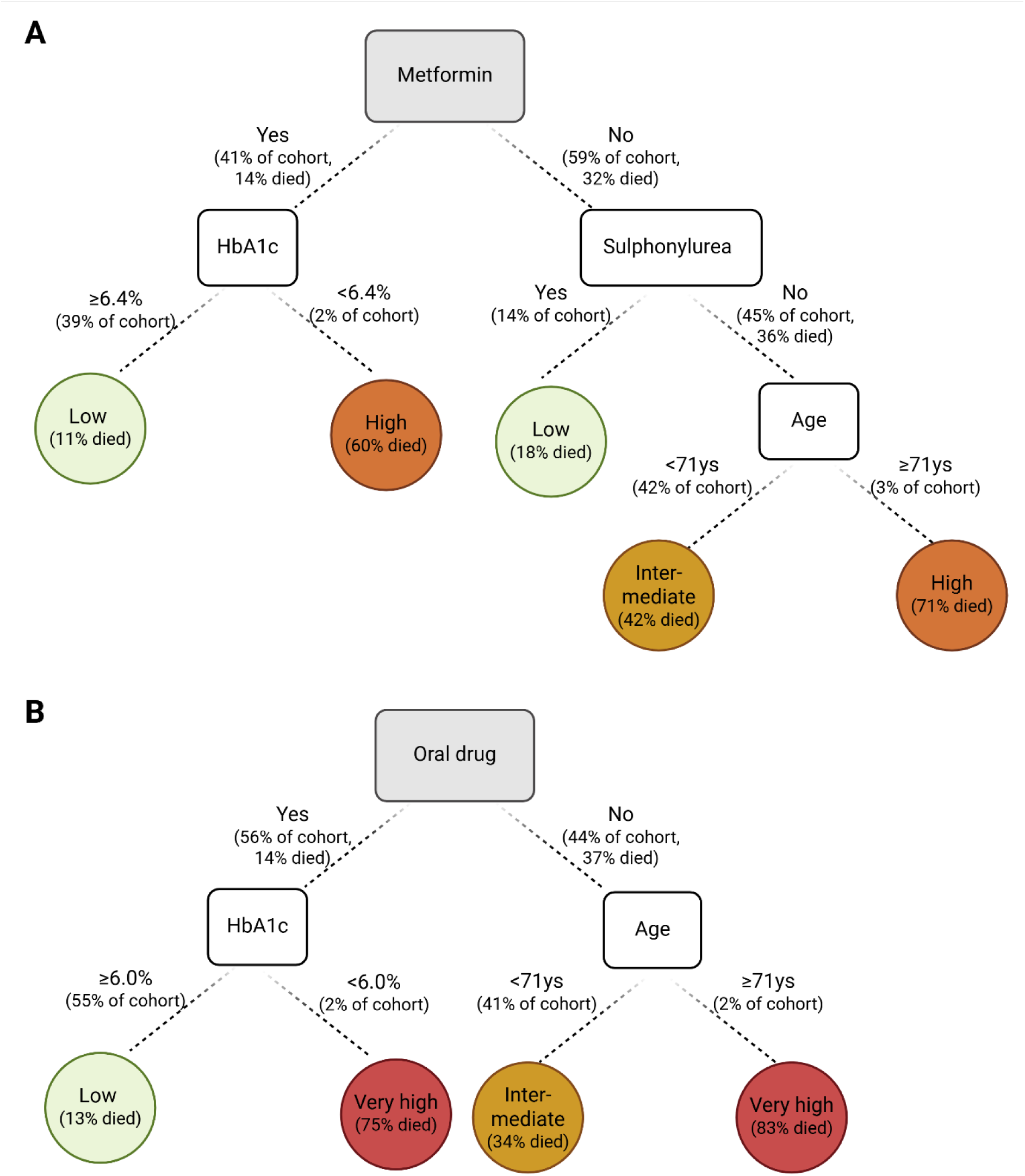
Fitted Classification Tree models of significant predictors of 28-day mortality among patients with acute melioidosis. Two modelling scenarios were used: (A) assuming separate oral drug (metformin and sulphonylurea use) included as distinct binary predictors and (B) assuming combined binary predictor to represent use of either metformin or sulphonylurea, along with adjusting for other covariates (sex, age, renal failure status, HbA1c, and insulin use). Coloured circles represent 28-day mortality rates in groups of individuals with various characteristics based on the classification tree decision points.

We next assessed model performance using a validation dataset and show varying levels of predictive accuracy and discriminative ability across the fitted logistic regression and classification tree models (**Figure 3**). The logistic regression model incorporating separate predictors for metformin and sulphonylurea achieved a prediction accuracy of 76.9% and an AUC of 0.754, indicating good overall model fit and good discriminative power. In contrast, the logistic model employing a combined oral drug indicator showed a higher prediction accuracy of 80.8%, though with a slightly lower AUC of 0.688, implying potential trade-offs between classification performance and discrimination. Similarly, the classification tree model that included metformin and sulphonylurea as separate variables demonstrated strong performance with an accuracy of 80.8% and an AUC of 0.767, whereas the tree model using the combined oral drug indicator had a slightly lower accuracy of 76.9% and an AUC of 0.658.

**Figure 3.**
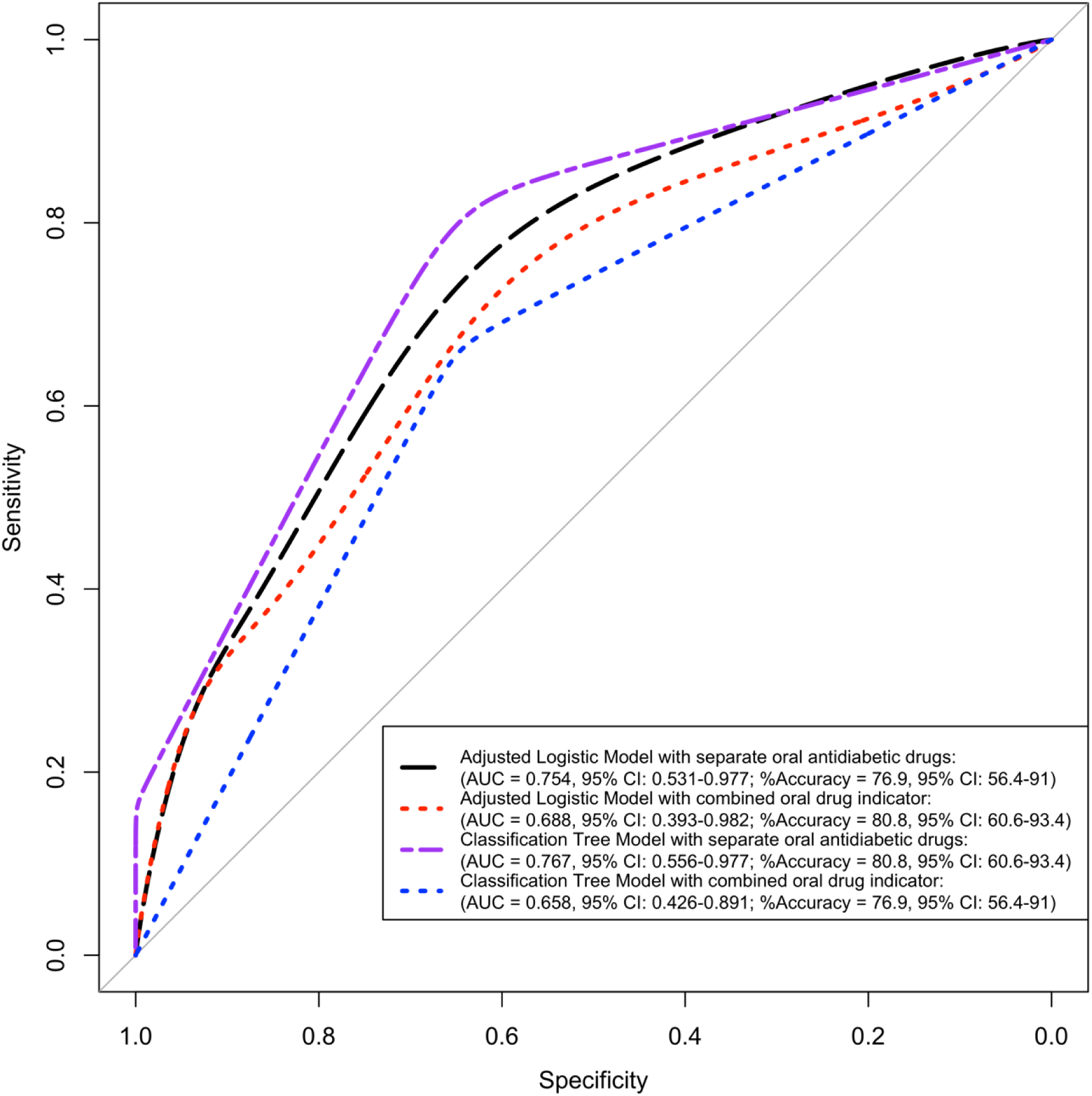
Predictive modelling performance comparing logistic regression and classification tree models. Comparative visualisation of the ROC curve of the best-fitted logistic regression and classification tree models under the two modelling scenarios with separate or combined oral anti-hyperglycaemic drugs.

### Favourable immune parameters in patients on metformin

For a subset of individuals, immunological data including antibody and T-cell responses to the bacteria[17] as well as targeted proteomics data were available. Since adaptive immune responses were highly variable between individuals and some patients did not mount any T cell or antibody responses at all (**Supplementary** Figure 1), we wanted to understand what drives immune responsiveness in this cohort. We interrogated the association of being a T cell and antibody responder, defined as greater than 16 IFN-γ spot forming units/million PBMC and a titre greater than 40 respectively, with demographic and clinical parameters in a multivariable logistic regression model. We found that individuals taking metformin had 3.5 fold increased odds of mounting a T cell response to BP antigens after adjusting for sex, age, HbA1c, 28-day mortality, history of renal impairment, insulin and sulphonylurea use (adjusted OR 3.52, 95% CI 1.15, 10.80, p=0.028), whereas mortality was associated with more than 70% decreased odds of mounting antibody responses to BP (adjusted OR 0.29, 95% CI 0.11, 0.78, p=0.014). Next, we assessed the effect of treatment on the expression of proteins involved in immune responses to infection in serum of 30 individuals. Permutation-based Hotelling’s T2 test revealed a significant global difference in cytokine profiles between Metformin-treated and untreated participants (Test statistic = –1.0745; permutation p = 0.0077). Using penalised multivariate analysis adjusting for age, sex, HbA1c, renal impairment, 28-day mortality, and sulphonylurea we identified 10 out of 44 cytokines whose expression levels were significantly associated with Metformin treatment (**Figure 4**, **Supplementary** Figure 2) with proteins involved in anti-bacterial defence (IL-12p70, IL-27, IL1RA, MIP-1b), T cell homeostasis (IL-7), angiogenesis and tissue repair (VEGFA+D, Flt1, Tie-2) being higher in the metformin group. Notably, the biggest treatment effect of metformin was seen for IL-12p70.

**Figure 4.**
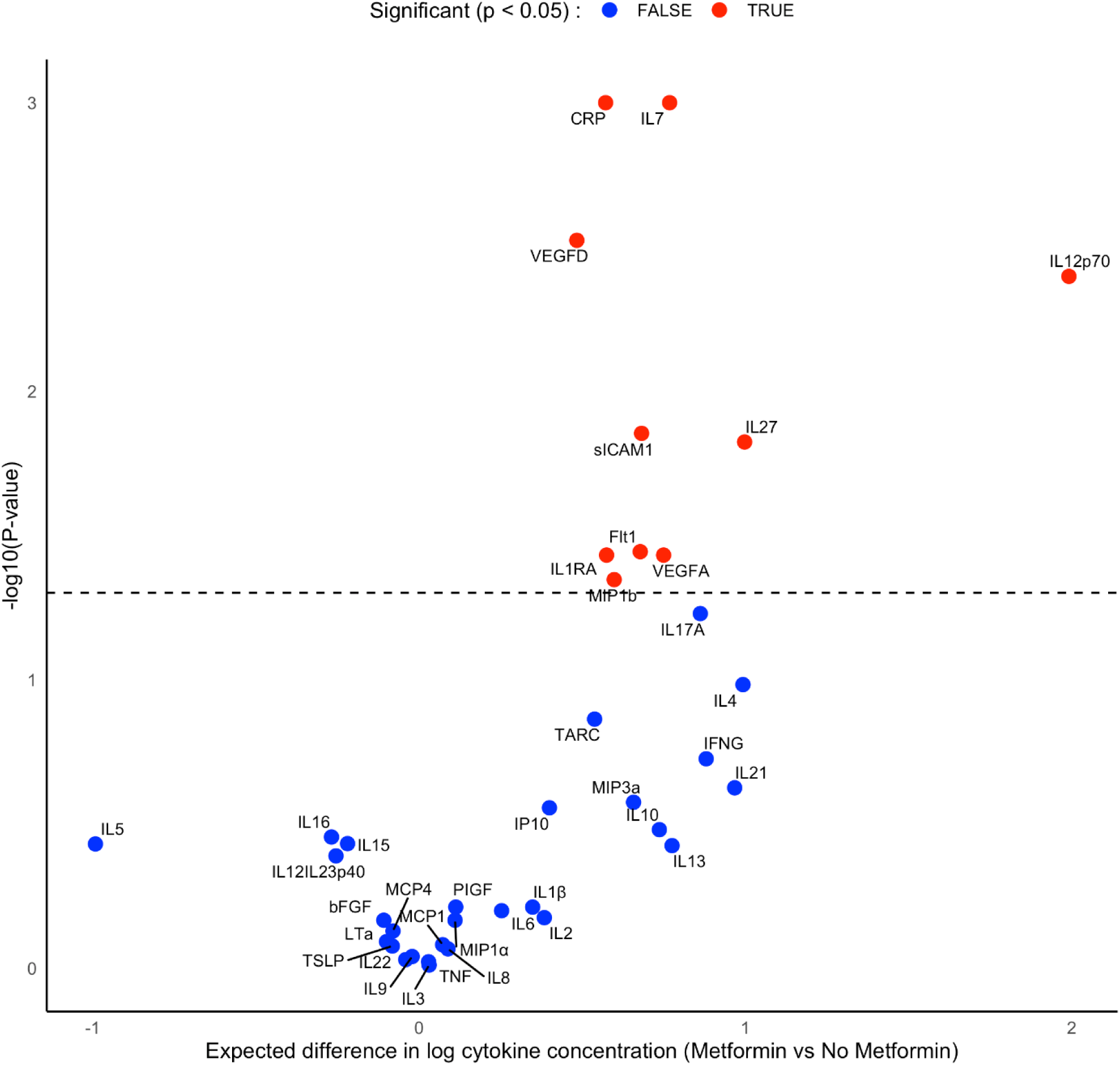
Volcano plot of cytokine expression changes between Metformin-treated and those with other treatments (sulphonylurea, insulin) derived from the Lasso-regularised multivariate regression model, adjusted for age, sex, HbA1c, renal impairment, 28-day mortality, and sulphonylurea use. Red points indicate cytokines with statistically significant changes (p<0.05); blue points indicate cytokines without significant changes (p≥0.05). P-values were obtained via a permutation-based procedure.

## DISCUSSION

Here we show that people with DM who have been taking metformin prior to infection are less likely to die from acute melioidosis, even after adjusting for use of sulphonylurea drugs, sex, renal impairment and HbA1c. This is in line with reports for COVID-19 [10] and TB [9]. Unsupervised classification tree analysis further highlighted the nuanced relationship between oral anti-hyperglycaemic drug use and mortality risk in this cohort, identifying glycaemic control (HbA1c) and age as modifying factors of outcome. Lower HbA1c in people with DM may be associated with increased frailty and weight loss from comorbid conditions such as cancer and malnutrition that can contribute to mortality [21]. Older individuals (≥71 years) not taking any oral drugs were at highest risk of death, reinforcing vulnerability of elderly patients. Furthermore, pre-existing chronic kidney disease was associated with worse outcome which is in line with a previous study on all-cause mortality in a Thai diabetic registry cohort [16].

There is insufficient data on the exact mechanisms of metformin action on anti-microbial responses in the context of DM and infection. Potential mechanisms include a direct anti-microbial effect, which has been described for several pathogens [22], the augmentation of autophagy and a reduction of inflammation and oxidative stress [6]. Metformin lowers hepatic glucose through substrate specific inhibition of gluconeogenesis (from lactate and glycerol) [23]. This glucose lowering action in itself might have a beneficial effect on disease tolerance. Metabolic changes during sickness are associated with a shift towards a catabolic state [24]. Importantly, glucose-restrictive conditions have been shown to increase survival in mouse models of bacterial sepsis due to a metabolic shift towards lipid utilisation and production of ketones [25]. We have previously shown that mortality is associated with decreased T cell responses to the pathogen [18]. Here we found that metformin treatment was associated with increased odds of mounting T cell responses to bacterial antigens, after adjusting for covariates including mortality. Aligned with this, levels of cytokines involved in T cell survival and homeostatic control of metabolism (IL-7) [26] and anti-bacterial response, specifically priming of T helper 1 responses (IL-12p70) [27] were elevated in the patient group taking metformin compared to those who did not. The soluble form of Tie-2, the endothelial cell specific tyrosine kinase receptor for angiopoietin (Ang) 2, was also elevated in the metformin group. Cleavage of Tie-2 has been shown to prevent Ang/Tie-2 signalling and soluble Tie-2 further functions as a trap for Ang-2 thereby potentially reducing its negative effect in sepsis [28]. Overall, this suggests a more balanced cellular response to infection in the metformin group.

Despite significant overlap in use of metformin and sulphonylurea drugs, the statistical models taking both into account as separate variables showed highest model performance. Nevertheless, a combined oral drug indicator clearly highlights the benefits of any oral drug treatment on outcome from melioidosis. Although the model taking separate oral drugs into account has better predictability, the simplified use of a single oral drug indicator still maintained clear predictive structure, making the model more concise and potentially more interpretable in clinical settings. A previous study conducted in the same region of Thailand demonstrated that sulphonylurea, specifically glyburide, was associated with better survival and this was associated with a reduced pro-inflammatory gene signature [15]. The study by Kho et al [15] was enriched for patients taking sulphonylurea with just over half of the DM patients taking glyburide and only 12% taking metformin, whereas our cohort was enriched for patients taking metformin (42%) and more closely representing the overall proportion of DM patients taking metformin [16]. Furthermore, the majority of patients in our study were on glipizide. While glyburide has previously been shown to have anti-inflammatory properties[15], it is unclear whether the same is true for glypizide. Although metformin is recommended as first-line therapy by the WHO [3] the proportion of patients taking metformin in our study was lower than what has been found in a large 3-year hospital-based national diabetes study in Thailand (6,373 out of 8,867 patients with T2DM taking metformin, 72%)[16]. Reasons for lower uptake are complex and include reduced tolerability of metformin in some patients [5]. In addition, availability and affordability of metformin varies greatly across the world and is lowest in low-income countries [29,30]. Combined evidence provided by this and other studies should be used to push policies on starting and maintaining metformin in everyone, if at all possible, and will require solutions to tackle drug tolerance and adherence issues. Exploring tolerance and cost-effectiveness of slow-release metformin alongside behavioural interventions to maintain drug adherence could be considered.

In conclusion, this study adds important evidence in favour of the protective effect of metformin. We also demonstrate that machine learning provides valuable information in addition to logistic regression and highlights predictors with a non-linear effect on outcome. Our results underscore the relevance of evaluating predictive models from multiple perspectives and highlight that a machine learning approach can further help inform clinical decisions in patients with acute melioidosis and diabetes. Future studies including randomised controlled trials and mechanistic studies are needed to causally link metformin with protection from infectious diseases.

### Limitations

There are a few limitations to this study. Firstly, information on duration of DM, length of treatment and previous treatment was unavailable. Secondly, confounding by indication is a potential bias in this study, whereby people not taking metformin may represent those with differences in overall health and / or less recently diagnosed, thus having had diabetes for a longer time. However, there was no difference in the proportion of people with renal impairment, a marker of complications of longstanding DM. Lastly, typing of DM was not available but the majority of DM in Thailand is thought to be Type 2 [31]. The presence of some young patients aged 19 years and up in the study population indicates further study of DM type and risk factors in the region is desirable.

## Acknowledgements

The authors would like to thank all the study participants, their families as well as the staff at Sunpasitthiprasong Hospital, Ubon Ratchatani.

## Author contributions

Conceptualisation: BK, SJD

Data Curation: BK, PA, PB, AW, KA

Formal Analysis: BK, CT, PA

Funding Acquisition: SJD, CC

Investigation: BK, CT, PA, SC, PC

Methodology: BK, CT

Resources: SJD, CC, NPJD

Supervision: BK, SJD, CC

Visualisation: BK, CT, PA

Writing-Original Draft Preparation: BK, CT

Writing-Reviewing&Editing: BK, CT, PA, SC, PC, PB, AW, KA, NC, DL, NPJD, PC, CC, SJD

## Data availability statement

Data is available from the corresponding authors upon reasonable request.

