## Supplementary Information for "Metformin use is associated with lower mortality from bacterial sepsis and improved immunocompetence in Thai diabetes patients with acute melioidosis"

Running title: Metformin is protective in Melioidosis

Barbara Kronsteiner[1,2], Clement Twumasi[1,3], Priyanka Abraham[1,2,4], Suchintana Chumseng[4], Panjaporn Chaichana[4], Phumrapee Boonklang[4], Arin Wongprommoon[4], Kesorn Angchagun[4], Narisara Chantratita[4,5], Direk Limmathurotsakul[4,6,7], Nicholas P.J. Day[4,7], Parinya Chamnan[8], Claire Chewapreecha[4,7], Susanna J. Dunachie[1,2,3,4,5]

1. Peter Medawar Building for Pathogen Research, Nuffield Dept. of Medicine, University of Oxford, Oxford, UK
2. NDM Centre for Global Health Research, Nuffield Dept. of Medicine, University of Oxford, Oxford, UK
3. NIHR Oxford Biomedical Research Centre, Oxford University Hospitals NHS Foundation Trust, Oxford, UK
4. Mahidol Oxford Tropical Medicine Research Unit, Faculty of Tropical Medicine, Mahidol University, Bangkok, Thailand
5. Department of Microbiology and Immunology, Faculty of Tropical Medicine, Mahidol University, Bangkok, Thailand
6. Department of Tropical Hygiene, Faculty of Tropical Medicine, Mahidol University, Bangkok, Thailand
7. Centre for Tropical Medicine and Global Health, Nuffield Department of Medicine, University of Oxford, UK
8. Cardiometabolic Research Group, Sunpasitthiprasong Hospital, Ubon Ratchathani, Thailand

### Methods

#### Serum protein data analysis (MSD)

Analysis of protein concentration in serum was performed using MSD Workbench 4.0 software, which generates a standard curve from the calibrators to determine the concentration of the marker that was multiplied by the dilution factor. An arbitrary value of 0.001 was added to overcome the presence of null values from samples below the detection range. Samples within detection range but below the standard curve were assigned a value that was half the concentration of the lowest standard for the analyte. For eight of the analytes at least 30% of the samples were below the fit curve range and were therefore excluded from analysis.

| **Supplementary Table 1. Analytes measured by MSD in serum of melioidosis patients.** | |
|---|---|
| Pro-inflammatory Panel 1 | IFN-γ, IL-1β, IL-2, IL-4, IL-6, IL-8, IL-10, IL-12p70, IL-13, TNF |
| Cytokine Panel 1 | GM-CSF, IL-1α, IL-5, IL-7, IL-12/IL-23p40, IL-15, IL-16, IL-17A, TNF-β, VEGF-A |
| Cytokine Panel 2 | IL-1RA, IL-3, IL-9, IL-17A/F, IL-17B, IL-17C, IL-17D, TSLP |
| Chemokine Panel 1 | Eotaxin, Eotaxin-3, IL-8, IP-10, MCP-1, MCP-4, MDC, MIP-1α, MIP-1β, TARC |
| Angiogenesis Panel 1 | FGF (basic), PlGF, Tie-2, VEGF-A, VEGF-C, VEGF-D, VEGFR-1/Flt-1 |
| Vascular Injury Panel 2 | CRP, ICAM-1, SAA, VCAM-1 |
| TH17 Panel 1 | IL-17A, IL-21, IL-22, IL-23, IL-27, IL-31, MIP-3α |

#### Statistical Analyses

The dataset, comprising 278 patients with a previous DM diagnosis prior to hospital admission, was examined for missing values, specifically due to five missing data points in the HbA1c variable. Little’s test for Missing Completely at Random (MCAR), implemented using the “naniar” R package, confirmed that the missingness was statistically MCAR. As a result, complete-case analysis was conducted using the remaining 273 patients. Two scenarios were taken into account for the statistical modelling: 1. *Separate Oral Drug Models:* —metformin and sulphonylurea use were included as distinct binary predictors and 2. *Combined Oral Drug Models*, where these two variables were replaced with a single binary indicator, Drug Oral, defined as “Yes” if the patient received either Metformin or Sulphonylurea (or both), and “No” otherwise. All models included additional covariates: sex, age, history of renal impairment, HbA1c (%), and insulin use. The dataset was randomly partitioned into a 90% training set (n = 247) for model fitting and a 10% testing set (n = 26) for cross-validation. Predictive performance was evaluated using the resulting confusion matrices and quantified by both prediction accuracy and the Area Under the Receiver Operating Characteristic Curve (AUC).

**Multivariate Comparison of Cytokine Profiles Between Treatment Groups**

To evaluate the overall effect of Metformin treatment on the high-dimensional cytokine profiles for the sub-cohort population, we first applied a robust permutation-based Hotelling’s T2 test. Given the relatively small sample size (n = 30) and the high-dimensional nature of the cytokine panel (with 44 outcomes), we employed the James-Stein shrinkage estimator for the sample covariance matrices. This approach mitigates issues arising from the non-normality of multivariate cytokine distributions (Royston test for multivariate normality, $p<0.001$), the small sample size relative to the number of outcomes, and high pairwise correlation between cytokines, without requiring prior dimension reduction.

The multidimensional cytokine profiles were found to be statistically inappropriate for principal component analysis (PCA) or dimension reduction based on the estimated Kaiser-Meyer-Olkin (KMO) statistic of 0.5 (although Bartlett’s Test of Sphericity was significant, $p<0.001$). Cytokine concentrations were log-transformed prior to analysis to stabilise variance and normalise distributions. Pairwise correlation matrices stratified by Metformin treatment and without stratification indicated strong correlations among cytokine outcomes within each group (**Supplementary Figures 3-5**).

**Lasso-Regularised Multivariate Regression to Estimate Adjusted Treatment Effects on Cytokine Profile**

To identify specific cytokines associated with Metformin treatment while accounting for potential confounders, we performed Lasso-regularised multivariate regression. Covariates or confounders including age, sex, HbA1c, renal impairment, 28-day mortality, and Sulphonylurea use were included in the model. The Metformin treatment variable was left unpenalised to preserve unbiased estimates of treatment effects, while penalisation was applied to covariates to reduce multicollinearity and eliminate uninformative predictors. Because penalised regression does not follow standard null distributions, p-values for the treatment coefficients were obtained via a permutation-based procedure such that: treatment labels were randomly shuffled, the penalised model was refitted while retaining Metformin unpenalised, and observed estimated regression coefficients were compared to the null distribution derived from over 1,000 permutations. The proportion of times each penalised covariate was selected across cytokine outcomes was evaluated, highlighting the most influential covariates or confounders and confirming that the estimated Metformin treatment effects remained robust to adjustment (**Supplementary Figure 6**).

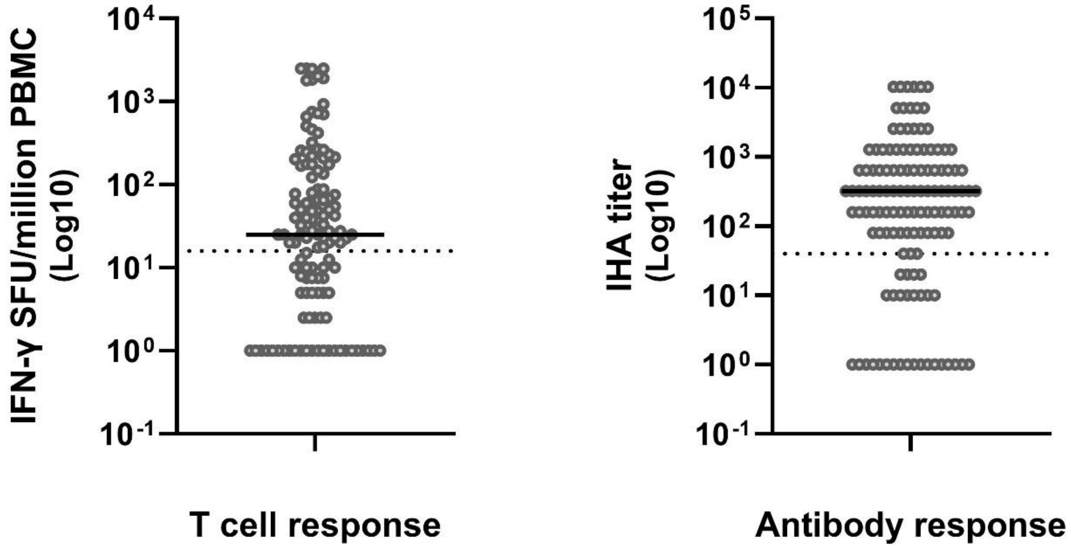


**Supplementary Figure 1.** T cell and antibody responses to *B. pseudomallei* were measured in peripheral blood mononuclear cells and serum of acute melioidosis patients. Dotted lines indicate the positivity cut-off above which an individual is considered a T cell or antibody responder, i.e. >16 SFU/million PBMC for T cell responses and a titre of >40 for antibody responses.

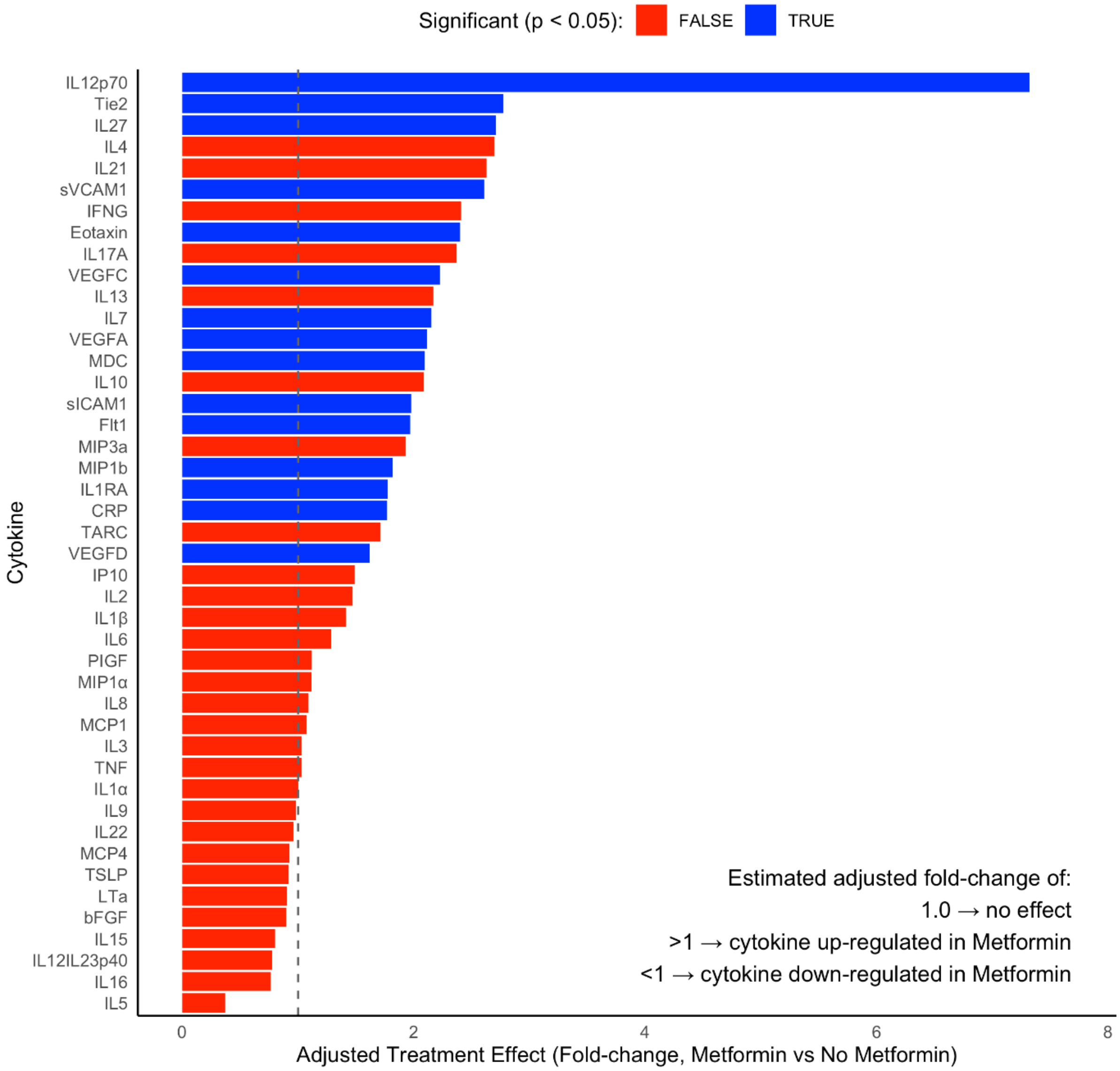


**Supplementary Figure 2.** Fold-change estimates of cytokine expression in the Metformin-treated group, derived from a Lasso-regularized multivariate regression model adjusted for age, sex, HbA1c, renal impairment, 28-day mortality, and sulphonylurea use. Fold-changes were calculated by exponentiating the model coefficients (values >1 indicate upregulation, <1 indicate downregulation, and 1 indicates no change). P-values were obtained via a permutation-based procedure.

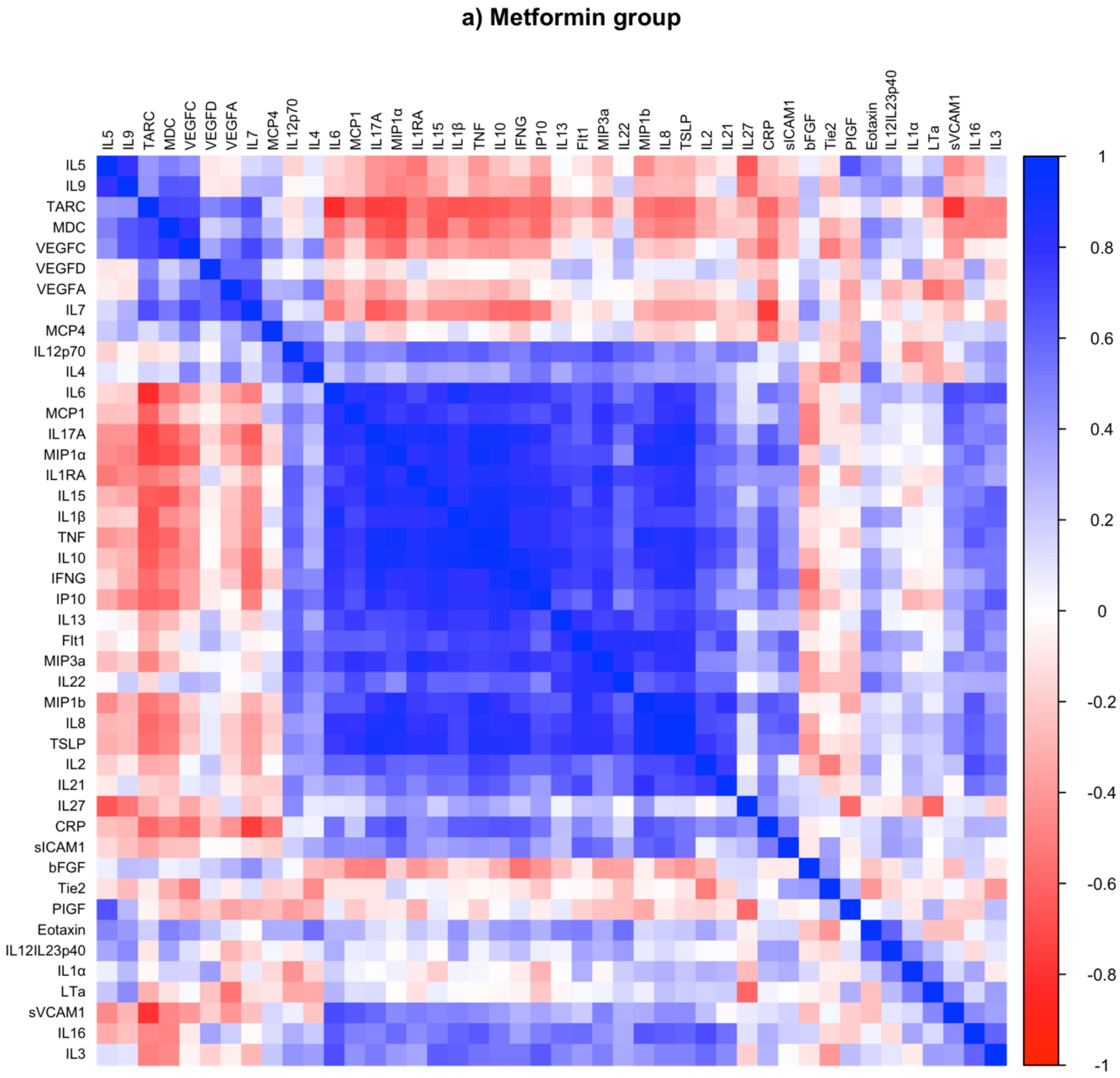


**Supplementary Figure 3**. Pairwise correlation matrix plot of the cytokine profiles for the Metformin sub-group.

### b) Non-Metformin group

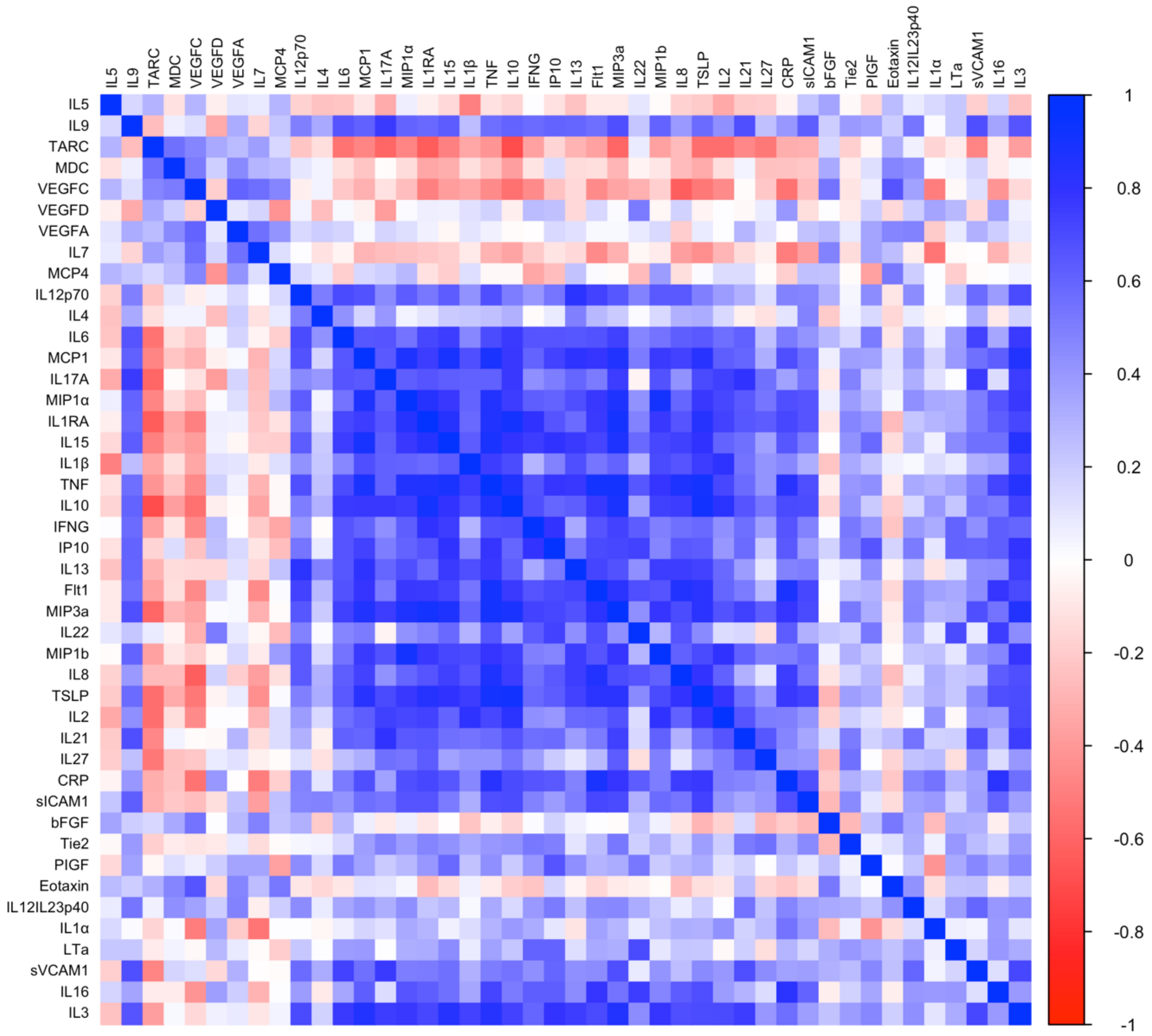


**Supplementary Figure 4.** Pairwise correlation matrix plot of the cytokine profiles for the non-Metformin sub-group

c) Combined treatment groups

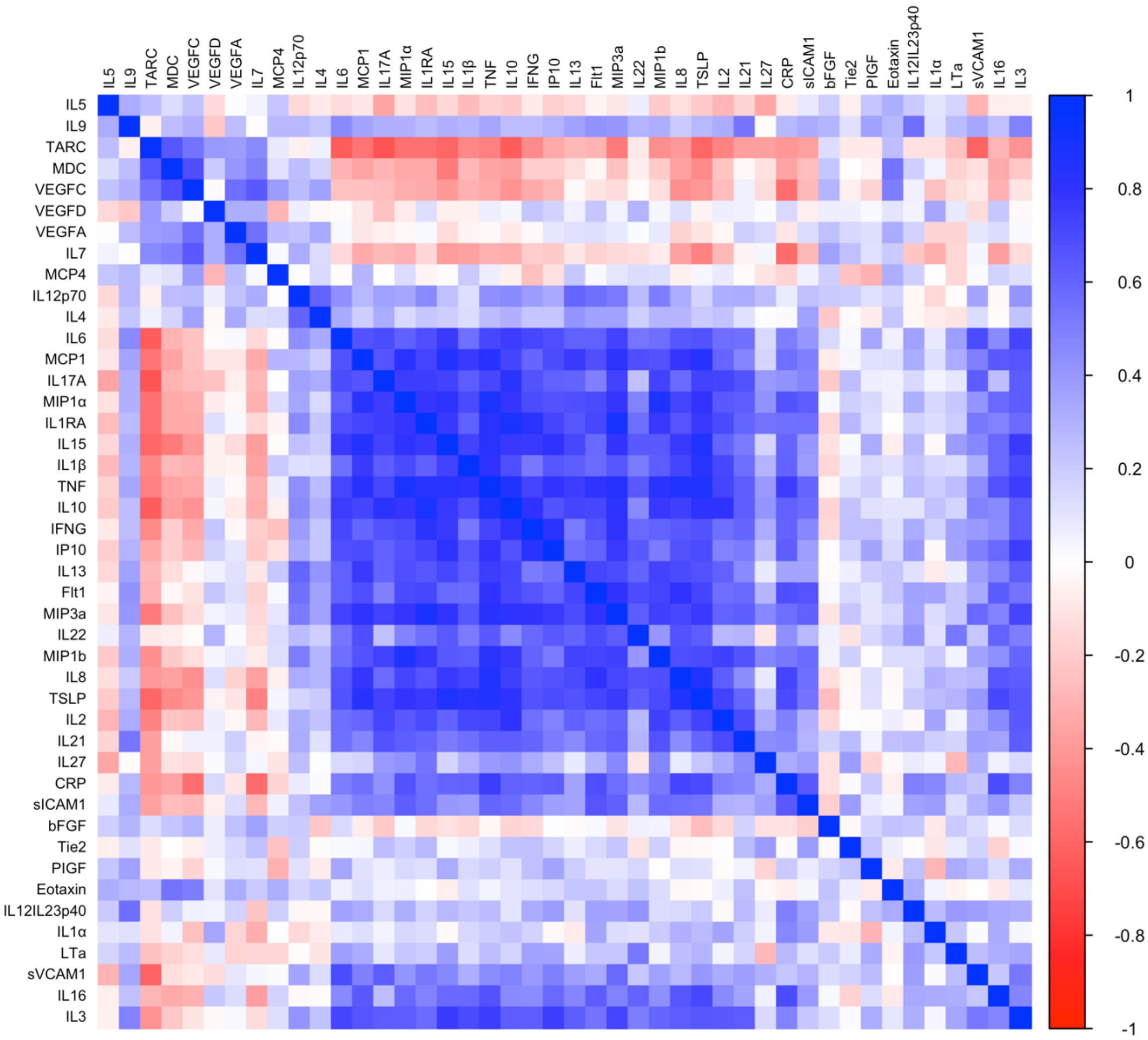


**Supplementary Figure 5.** Pairwise correlation matrix plot of the cytokine profiles without stratification by Metformin treatment groups.

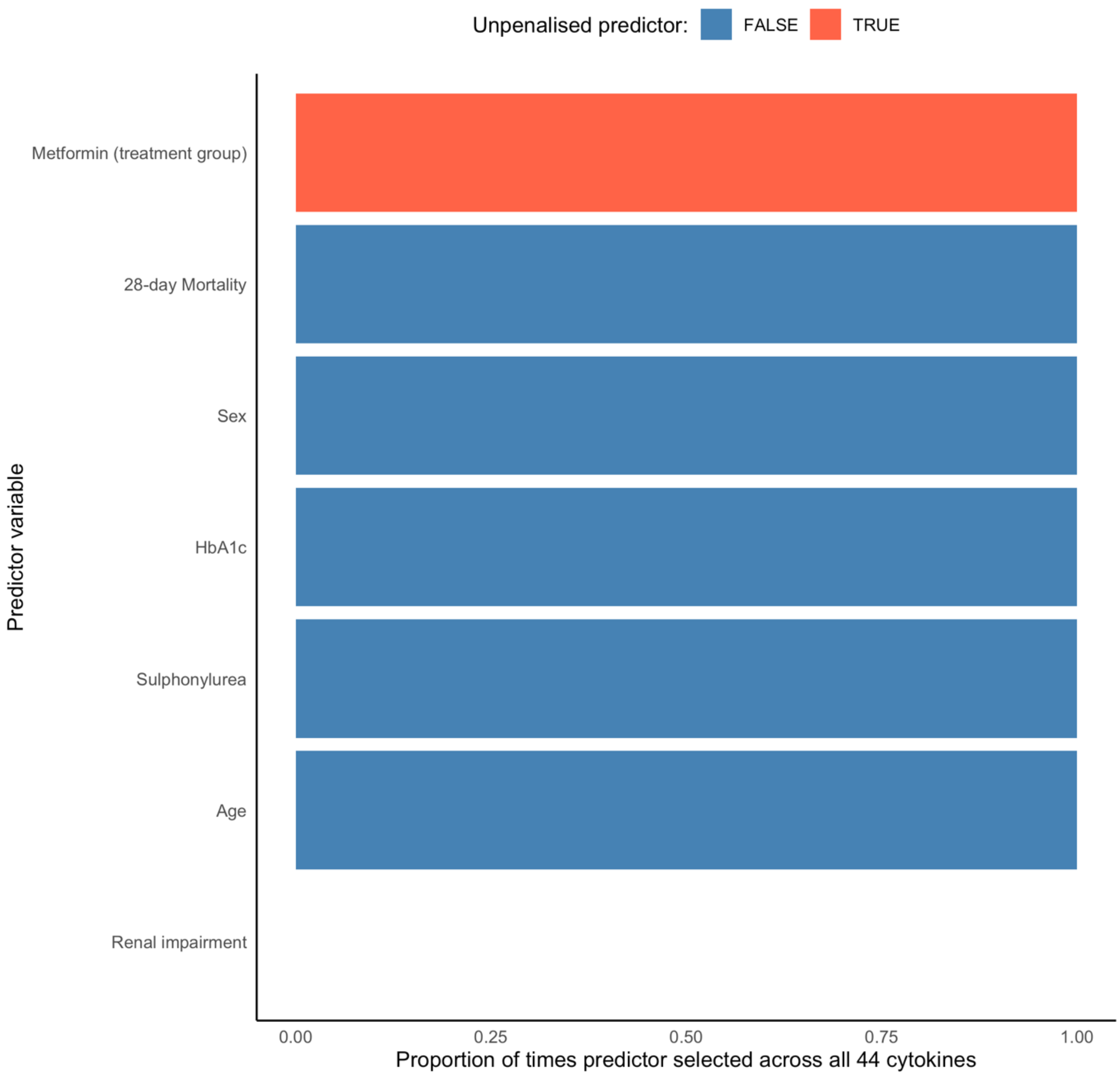


**Supplementary Figure 6.** Proportion of times each penalised covariate was selected across cytokine outcomes in the Lasso-regularised multivariate regression model. Covariates selected in a higher proportion of outcomes are shown as more influential, highlighting key confounders while demonstrating that the estimated Metformin treatment effects were robust to adjustment.
